# The burden of Progressive Fibrotic Interstitial lung disease across the UK

**DOI:** 10.1101/2020.11.16.20229591

**Authors:** Thomas Simpson, Shaney L Barratt, Paul Beirne, Nazia Chaudhuri, Anjali Crawshaw, Louise E Crowley, Sophie Fletcher, Michael A Gibbons, Philippa Hallchurch, Laura Horgan, Ieva Jakaityte, Thomas Lewis, Tom McLellan, Ryan Miller, Stefan Stanel, Muhunthan Thillai, Fiona Thompson, Zhe Wu, Philip L Molyneaux, Alex G West

## Abstract

While Idiopathic pulmonary fibrosis (IPF) remains the exemplar progressive fibrotic lung disease, there remains a cohort of non-IPF fibrotic lung diseases (fILD) which adopt a similar clinical behaviour to IPF despite therap. This phenotypically related group of conditions, where progression of disease is similar to that seen in IPF, have recently been described as Progressive Fibrotic Interstitial Lung diseases (PF-ILD). Previous estimates suggest that between 18 to 40% of all fILD will develop progressive disease, however, the exact burden remains unknown. This retrospective, observational study therefore aimed to estimate the incidence of PF-ILD across England.

All new referrals seen across nine UK centres for their first outpatient clinic appointment between 1st August 2017 and 31st January 2018 were assessed against the diagnostic criteria for PF-ILD laid out in the INBUILD trial. A total of 1749 patients with fILD were assessed. In this cohort of patients at risk of developing PF-ILD the INBUILD criteria were met in 14.5% (253/1749) of all new non-IPF fILD referrals. The average time from referral to specialist centre to diagnosis of progressive phenotype was 311 days. Of the progression events the majority were driven by a measured drop in FVC, with more than half of patients experiencing a drop of 10%. Almost one quarter of patients (24.1%) were diagnosed with progressive disease on the basis of radiological and symptomatic progression alone without a spirometric deterioration.

This study represents a fair and balanced approach to assessing the incidence of objectively measurable and treatable PF-ILDs in the UK. A rate of 14.5% of new referrals with non-IPF ILD is less than that reported in previous studies however our methodology is likely to give a more accurate result than estimates based on extrapolation from general disease statistics, from physician-reported estimates prone to significant biases, or insurance claim processes also substantially prone to bias. This information has implication for workforce planning and the funding of anti-fibrotic therapy in the UK and beyond.

## To the Editor

While Idiopathic pulmonary fibrosis (IPF) remains the exemplar progressive fibrotic lung disease, there remains a cohort of non-IPF fibrotic lung diseases (fILD) which adopt a similar clinical behaviour to IPF despite therapy [1]. This phenotypically related group of conditions, where progression of disease is similar to that seen in IPF, have recently been described as Progressive Fibrotic Interstitial Lung diseases (PF-ILD) [2]. Historically treatments for these cases have been limited and clinicians, recognising the related progression between these conditions and IPF may have been pragmatically relabelling these cases as IPF based on their disease behaviour to qualify for anti-fibrotic therapy [3]. The INBUILD trial broadened the scope of treatable fILD by demonstrating a significant benefit of Nintedanib in patients with fILD and progressive disease [4]. In response to this the European Commission (EC) approved an additional indication for Nintedanib in adults for the treatment of PF-ILD in July 2020.

While research interest grows in the progressive phenotype and debates about the optimal diagnostic criteria continue the incidence of patients with PF-ILD potentially eligible for treatment according to the criteria laid out in the INBUILD trial remains unclear. Previous attempts to estimate the proportion of fILD who develop a progressive fibrotic phenotype have either used estimates based on the disease behaviour of individual conditions [5], interviews with experts [6] or analysis of insurance claims [7]. This has resulted in estimates ranging from 18 to 40% of all fILD that will develop progressive disease. With the anticipated approval of therapeutic interventions for this cohort of patients worldwide, including in the UK, there is an urgent need to refine these estimates in a real-world population to enable appropriate service provision.

This retrospective, observational study therefore aimed to estimate the incidence of PF-ILD across England. Nine centres providing commissioned tertiary referral services for ILD were included. All new referrals seen for their first outpatient clinic appointment between 1st August 2017 and 31st January 2018 were assessed against the diagnostic criteria for PF-ILD laid out in the INBUILD trial [8] and in particular, the criteria for progression: relative decline in FVC % predicted ≥10%, or FVC decline ≥5% but <10%, combined with worsening respiratory symptoms, or FVC decline ≥5% but <10%, combined with radiological progression; or radiological progression with worsening respiratory symptoms. Continuous variables are presented as means (± Standard Deviation [SD]), and categorical variables as proportions.

A total of 2368 patients with fILD were assessed across the 9 centres. Six hundred and nineteen patients were diagnosed and managed as IPF and therefore excluded, leaving 1749 patients with fILD who were screened against the INBUILD criteria for progression, to identify cases of PF-ILD either at the first clinical review, or in the subsequent 2 years of follow up (Table 1). In the cohort of patients at risk of developing PF-ILD the INBUILD criteria were met in 14.5% (253/1749) of all new non-IPF fILD referrals, with a range between these specialist ILD centres from 8.9% to 23.6% of total cases. The average time from referral to specialist centre to diagnosis of progressive phenotype was 311(±273) days.

**Table 1:** Determinants of progressive disease.

| INBUILD CRITERIA | Number of cases meeting this criteria (% of PF-ILD cohort) |
| --- | --- |
| Relative FVC decline $\geq 10\%$ | 132 (52.2%) |
| FVC decline $\geq 5\%$ - $< 10\%$ with radiological progression | 40 (15.8%) |
| FVC decline $\geq 5\%$ - $< 10\%$ with symptom progression | 20 (7.9%) |
| Radiological and symptom progression | 61 (24.1%) |

The most common diagnoses associated with a PF-ILD phenotype were chronic hypersensitivity pneumonitis (84/253, 33.2%), unclassifiable ILD (44/253, 17.3%), connective tissue disease-associated ILDs including rheumatoid arthritis-associated ILD (42/253, 16.6%) and non-specific Interstitial pneumonitis (36/253, 14.2%). In the PF-ILD cases, the mean age was 68 ±12.4 years and interestingly 53.4% of the cohort was female, as compared to the well-recognised male predominance seen in IPF.

Of the progression events the majority were driven by a measured drop in FVC, with more than half of patients experiencing a drop of ≥10%. Almost one quarter of patients (24.1%) were diagnosed with progressive disease on the basis of radiological and symptomatic progression alone without a spirometric deterioration (Table 1).

The variations between centres and clinicians in diagnostic pathways, approaches to follow-up and definitions of progression has previously made it difficult to define and assess this cohort of patients. One of the strengths of our approach was the central collation and uniform application of the INBUILD inclusion criteria. However, this was done retrospectively and this is the main limitation of our study. While the INBUILD trial criteria are mostly objectively measurable phenomena, the definition of progressive symptoms may allow some biasing towards inclusion in those cases where spirometric progression was either not evidenced or not available, thus increasing the numbers of cases. Over a quarter of referrals received a final multidisciplinary team (MDT) diagnosis of IPF, and this is often pragmatic and based on their clinical disease behaviour, to allow access to antifibrotic therapy. However, a patient’s initial clinical and radiological features may have had more in keeping with a different ILD but with a PF-ILD phenotype. While we sought to ensure that all diagnostic labels in this study had been agreed at a local MDT, we did not at any point seek to reassess the accuracy of those labels and therefore some cases of non-IPF PF-ILD may have been missed. However, this burden of IPF is in keeping with historical data of the caseload of IPF across the enrolling centres and importantly reflects clinical practice, which we aimed to capture.

## Data Availability

Anonymised data is available on request

